# Preventable deaths involving opioids in England and Wales, 2013-2022: a systematic case series of coroners’ reports

**DOI:** 10.1101/2022.11.16.22282411

**Authors:** Francesco Dernie, Elizabeth T Thomas, Maja Bilip, Nicholas J. DeVito, Harrison S. France, Robin E. Ferner, Anthony R. Cox, Carl Heneghan, Jeffrey K. Aronson, Georgia C. Richards

## Abstract

**Background:** Deaths from opioids have increased in England and Wales, despite recognition of their harms. Coroners’ Prevention of Future Death reports (PFDs) provide important insights that may enable safer use and avert harms, yet these reports involving opioids have not been synthesised. We therefore aimed to identify opioid-related PFDs and explore concerns expressed by coroners to prevent future deaths.

**Methods:** In this systematic case series, we screened 3897 coronial PFDs dated between 01 July 2013 and 23 February 2022. These were obtained by web scraping the UK’s Courts and Tribunals Judiciary website to create an openly available database: https://preventabledeathstracker.net/. PFDs were included when an opioid was implicated in the death. Included PFDs were descriptively analysed, and content analysis was used to assess concerns reported by coroners and responses to such concerns.

**Findings:** Opioids were involved in 219 deaths reported by coroners in PFDs (5·6% of all PFDs), equating to 4418 years of life lost (median 33 years/person). Morphine (29%), methadone (23%), and diamorphine (16%) were the most common implicated opioids. Coroners most frequently raised concerns regarding systems and protocols (52%) or safety issues (15%). These concerns were most often addressed to NHS organisations (51%), but response rates were low overall (47%).

**Interpretation:** Opioids could be used more safely and appropriately if coroners’ concerns in PFDs were addressed by national organisations such as NHS bodies, government agencies, and policymakers, as well as individual prescribing clinicians.

**Funding:** No funding was obtained for this study. The National Institute for Health Research (NIHR) School for Primary Care Research (SCPR) provided funding to establish the Preventable Deaths Tracker website: https://preventabledeathstracker.net/

**Research in context:** *Evidence before this study:* We conducted a systematic search of PubMed and Google Scholar to identify studies of deaths involving opioids in England and Wales and analyses of coroners’ Prevention of Future Death reports (PFDs). We found that deaths from opioids had increased in England and Wales. We also identified studies that have used PFDs to assess preventable deaths during the COVID-19 pandemic and deaths involving anticoagulants, medicines purchased online, medication errors and adverse drug reactions. However, no study to date has examined opioid-related PFDs.

*Added value of this study:* We analysed coroners’ concerns in opioid-related deaths for which they issued PFDs and found that on average three decades of life are lost per individual. We found that most opioid-related PFDs involved males (64%) and were caused by prescribed opioids (52%). Deaths involving illicit opioids (24%) were more likely to occur in younger males than deaths from prescribed opioids. Failures in systems and processes were most commonly found to have contributed to preventable opioid-related deaths, but more than half of such concerns remain unaddressed.

*Implications of all the available evidence:* Coroners’ PFDs offer important real-world insights into opioid-related deaths and can inform public health strategies that aim to improve the safe use of opioids. Future work should focus on disseminating these findings more widely and engaging with key stakeholders such as NHS organisations and government agencies, so that findings from PFDs can inform guidelines and be implemented in clinical practice.

## Introduction

In 2019, the UK had the highest rate per capita of opioid consumption globally.^1^ Prescriptions of opioids in primary care and the rates of opioid-related hospitalisation in the UK have both increased contemporaneously^2-4^. In England and Wales, deaths from opioids have increased nearly five-fold in under three decades, from 8·4 per million of the population in 1993 to 39·7 per million of the population in 2020.^5^

In England and Wales, unnatural or unclear causes of deaths are established at inquests conducted by coroners. Since 1984, coroners have had a duty to report and communicate a death when they believe that action can be taken to prevent future deaths.^6^ These reports, named Prevent Future Deaths (PFDs) are now mandated by the Coroners and Justice Act 2009 and the Coroners (Investigations) Regulations 2013.^7,8^

Studies have systematically analysed PFDs to assess deaths during the COVID-19 pandemic, involving anticoagulants, medicines purchased online, medication errors and adverse drug reactions^9-13^. A case series of medicine-related PFDs showed that opioids were the most common drug involved in deaths.^14^ However, these concerns have not been synthesised. A systematic assessment of opioid-related PFDs and their associated responses might therefore provide valuable information to inform public health strategies to reduce opioid deaths.

The aim of our study was to systematically conduct a case series of PFDs to identify and characterise deaths involving opioids, synthesise coroners concerns and classify the responses of individuals or organisations to whom PFDs were addressed.

## Methods

A systematic case series was designed and the study protocol was preregistered on an open repository.^15^

### Data collection

Data were acquired from the Courts and Tribunals Judiciary website^16^ using web scraping^17^ to populate the Preventable Deaths Database^18^, which included 3897 PFDs as of 23 February 2022. The code that generates the database is openly available.^19^

### Data screening and eligibility

The 3897 PFDs were independently screened by study authors (GCR, MB, HF, FD) using a predefined definition for opioids; any substance, either natural, synthetic, or semi-synthetic, “derived from or having properties similar to those of morphine”.^20^ Cases were included if an opioid was believed to have caused or contributed to the death, including licit and illicit opioids, as per a pre-defined algorithm for case inclusion (Supplementary Figure S1).

### Data extraction

The web scraper automatically extracted the case reference number; date of the PFD report; name of the deceased; coroner’s name; coroner’s area; category of death (as defined by the Chief Coroner’s office); the institution(s) or individual(s) to whom the report was sent; and the URL of the case.

The investigators (FD, GCR, HSF, ETT) manually extracted the following variables into the database: the dates on which the inquest began and ended; number of deceased mentioned in the PFD; dates of death(s); sex; age; setting/location of death; any relevant medical and/or social history; medical cause(s) of death and/or conclusion of the coroner’s inquest; substance(s) implicated in deaths, including any reported opioids; details of the coroner’s concern(s); the number and types of individual(s) or organisation(s) to whom the reports were sent; whether the individuals or organisations addressed responded (yes/no), the date of the response, and the number of days under or overdue.

We also extracted mortality statistics for England and Wales on opioid-related deaths from the Office for National Statistics (ONS) from 2013 to the most up-to-date data (2020)^5^ at time of analysis (April 2022) to show the rates of opioid-related deaths.

### Data analysis

The number of included opioid-related PFDs, their rates as a proportion of all PFDs, and ONS mortality data were plotted over time. Medians and interquartile ranges (IQRs) were calculated for continuous variables (e.g. age) and frequencies were reported for categorical variables (e.g. sex, type and source of opioid, coroner area of jurisdiction).

We calculated the years of life lost (YLL) for the deceased with available ages using the formula, *YLL* = ∑(*E* – *A*); where *E*=life expectancy (WHO estimate of 75 years); and *A*=age of subject at death^21^. When age at death was over 75, these were described individually and counted as ‘0’ YLL.

One investigator (FD) assigned the International Statistical Classification of Diseases and Related Health Problems 10^th^ Revision (ICD-10)^22^ numeric codes for the causes of death to each PFD and compared the number of PFDs correctly categorised under the alcohol, drug and medication related deaths section on the Judiciary website.

To examine geographical variation, we graphed both the absolute counts and rates of opioid-related PFDs per all PFDs in each coroner area mapped to the standard regions of England and Wales.

To collate and evaluate concerns raised by coroners, we classified and identified repeated themes using directed content analysis.^23^ This allowed us to highlight cases involving previously recognised concerns, and to explore concepts, drawing similarities and disparities from the data.

To calculate response rates to PFDs, we used the 56-day legal requirement^7^ to classify responses as “early” (>7 days before due date), “on-time” (±7 days before due date), “late” (>7 days after due date), or “overdue” (response was not available on the Judiciary website as of 23 February 2022). We calculated the average response rate and frequency for individuals and organisations.

Available demographics and coroners’ concerns were also reported by the source of opioids (i.e. prescribed vs. illicit opioids) in a sub group analysis.

### Software and data sharing

We used Datawrapper^24^ to produce a choropleth map of England and Wales, and Tableau software to generate our bubble chart of coroners’ concerns. The study protocol, materials and statistical code are openly available via the Open Science Framework^15^ and GitHub.^19^

## Results

There were 219 opioid-related PFDs and deaths between July 2013 and 23 February 2022 in England and Wales (5·6% of all PFDs). The rate of opioid-related PFDs increased from 2.3% in 2013 to 7·3% in 2022 (Figure 1). Compared with ONS statistics, a median of 1·2% (IQR: 0·84-1·29) of all opioid-related deaths in England and Wales were written into PFDs each year (Supplementary Table S1).

**Figure 1.**
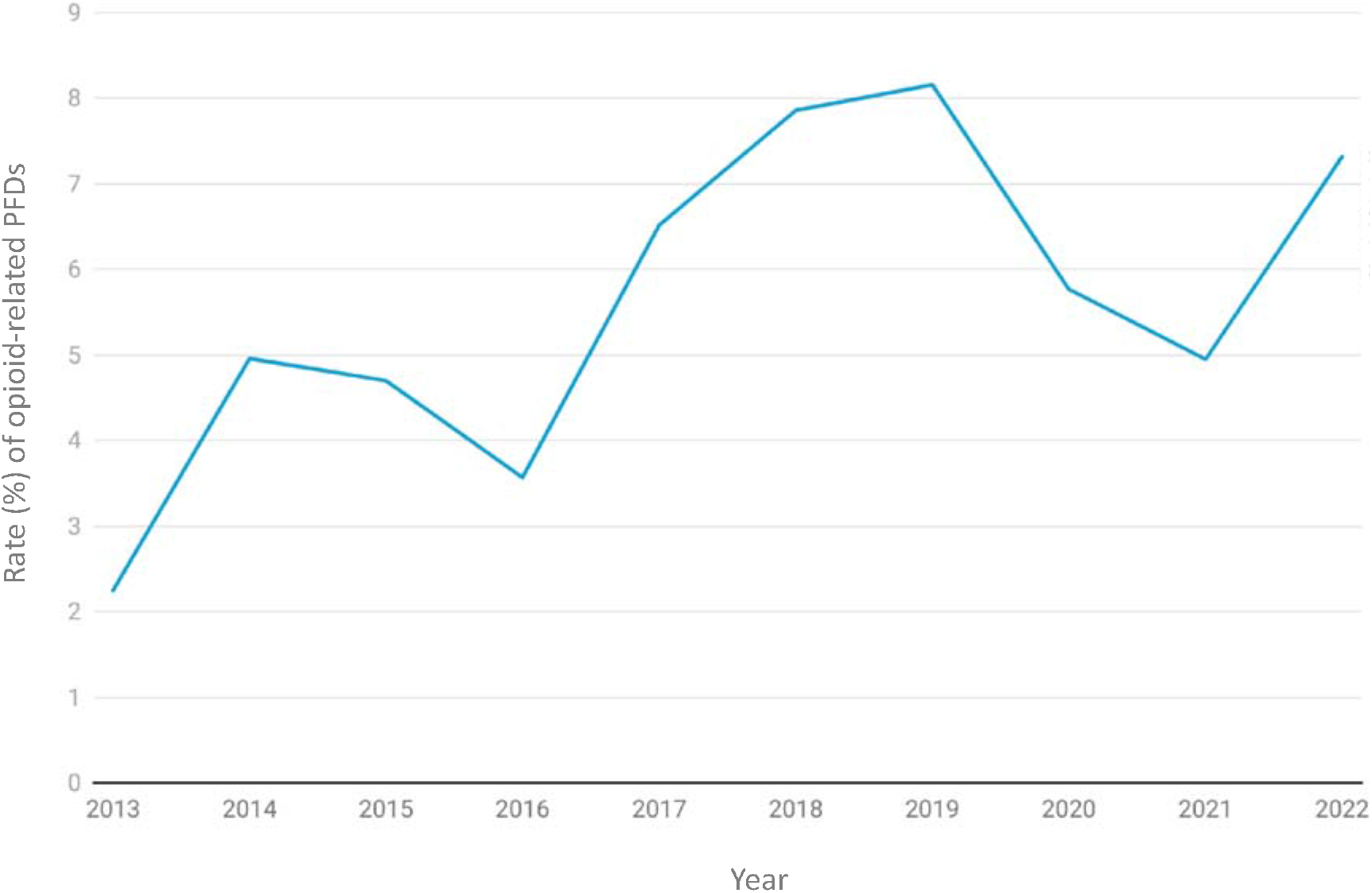
The rate of opioid-related Prevention of Future Deaths reports (PFDs) as a proportion of all PFDs (n=3897) published between 1 July 2013 and 23 February 2022 in England and Wales.

In these preventable deaths, 4418 years of life were lost, a median of 33 years per person (IQR: 24-42·5; n=137). The median age at death was 42 years old (IQR: 32·5-51 years; n=137) and five individuals were older than 75 years at the time of death. The average age at death was almost a decade younger in those that sourced illicit opioids (35 years; IQR: 30·3– 44; n=32) compared with prescribed opioids (43 years; IQR: 31-58·3; n= 70 Supplementary Table S2).

In all opioid-related PFDs, most (64%, n=141) of those who died were male. However, those that obtained illicit opioids had a higher proportion of males (86·5%; n=52) compared to the prescribed group (50%; n= 144; Supplementary Table S2).

On the Judiciary website, only 37% (n=82) of opioid-related PFDs were classified under the ‘Alcohol, drug and medication related deaths’ report type and 4% (n=8) were classified as ‘Product related deaths’ (Supplementary Table S3). However, when applying the ICD-10 categorisation, the most common (41%) causes of death were poisoning by ‘other’ opioids (e.g. morphine, codeine), followed by poisoning by methadone (20%) and poisoning by other synthetic narcotics (19%) (Supplementary Table S4).

Morphine (29%) was the most frequently implicated opioid in deaths, followed by methadone (23%), and heroin (16%) (Figure 2). Most (52%, n=114) of the deaths involved opioids that were prescribed, followed by illicit drugs (24%, n=52), and a combination of licit and illicit (14%, n=31). In 22 cases (10%) the source of the drug was unclear or unstated. Those for whom opioids were prescribed commonly received analgesics and methadone, compared with those who obtained illicit opioids, who most often sourced heroin (Supplementary Figure S2).

**Figure 2.**
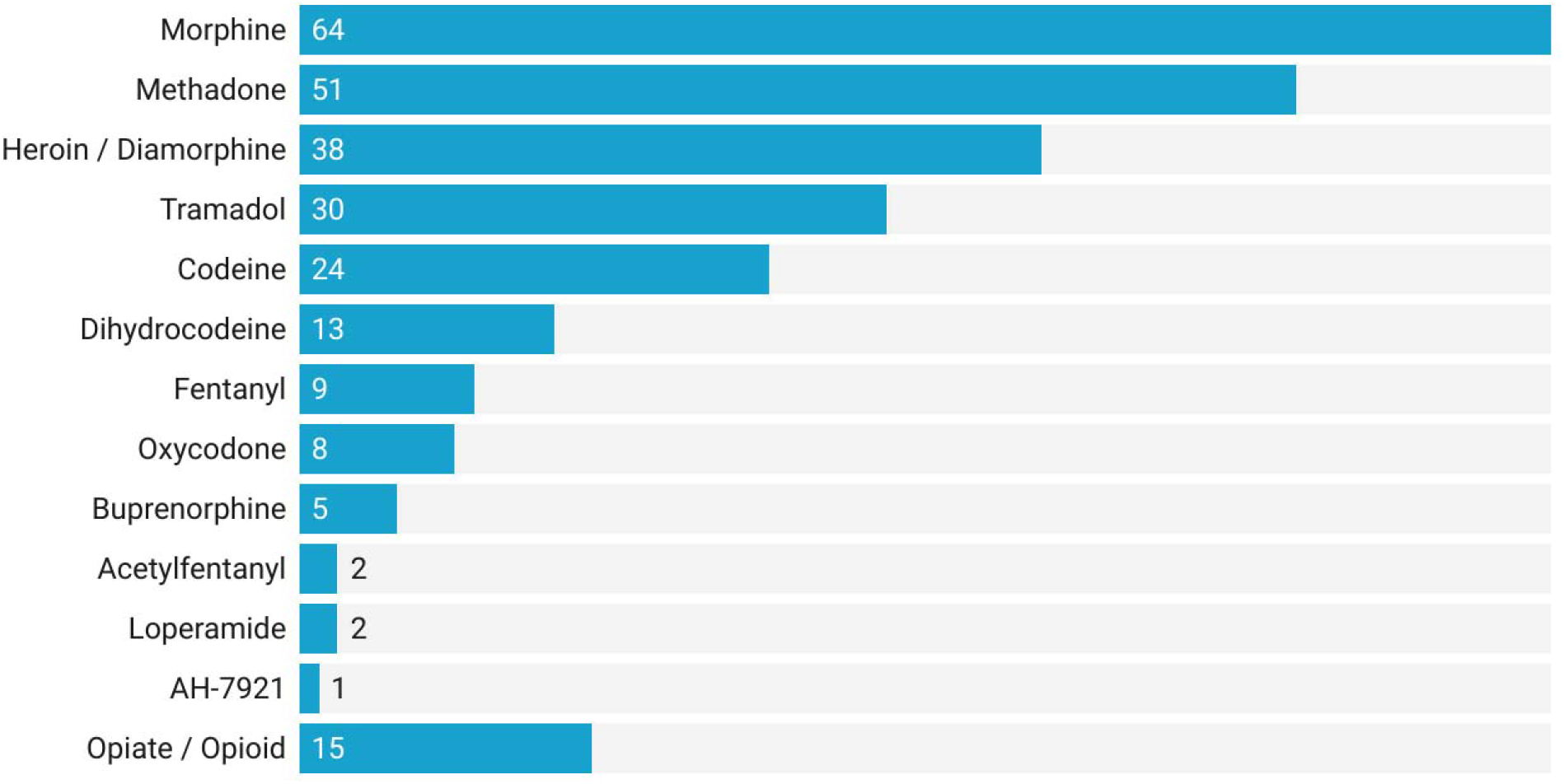
Rates of opioid drugs reported in 219 Prevent Future Death (PFD) reports published between July 2013 and February 2022 in England and Wales. Some PFDs reported multiple opioids and others did not specific the type of opioid involved (i.e. opiate/opioid category).

There were 219 PFDs, written by 125 coroners across 71 jurisdictions (Supplementary Table S5). Coroners in the North-West of England wrote the most (25%, n=55/219) opioid-related PFDs (Figure 3a); however, after correcting for the number of PFDs written in each area, the highest rate of opioid-related PFDs (18·5%) was reported in the South-West of England (Figure 3b).

**Figure 3.**
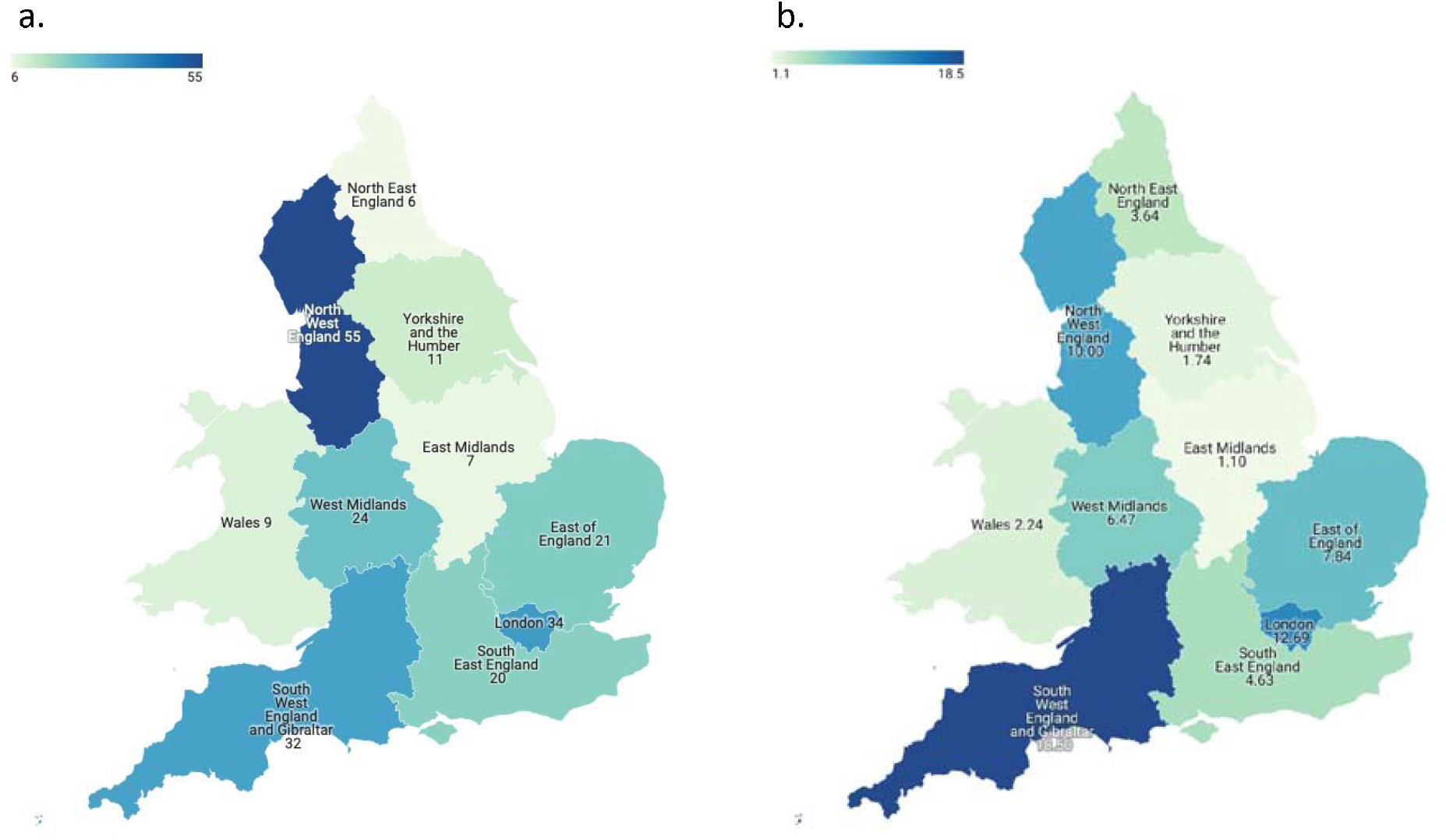
Choropleth map of the regions of England and Wales from which the 219 opioid-related Prevention of Future Deaths reported (PFDs) were issued by coroners between July 2013 and February 2022, showing the raw numbers of PFDs published (a), and the rate of opioid-related PFDs per all PFDs published (b).

Coroners expressed 666 individual concerns in the 219 opioid-related PFDs. Using content analysis, these concerns were categorised into 42 separate themes with five higher-order themes, which were systems and protocols, communication, safety, education and training, and resources. In all opioid-related PFDs, concerns were most often related to systems and protocols (52%, Supplementary Table S6), and this was reflected in both the illicit (51%) and prescribed (51%) groups (Supplementary Table S7). The most common individual concerns for all opioid-related PFDs were the failure to monitor/observe patients (10%), poor communication between organisations (8%), and unsafe protocols (7%; Figure 4; Supplementary Table S6). In people for whom opioids were prescribed, concerns of excessive supply and inappropriate dosing were more common than in those who used illicit opioids (Supplementary Table S7).

**Figure 4.**
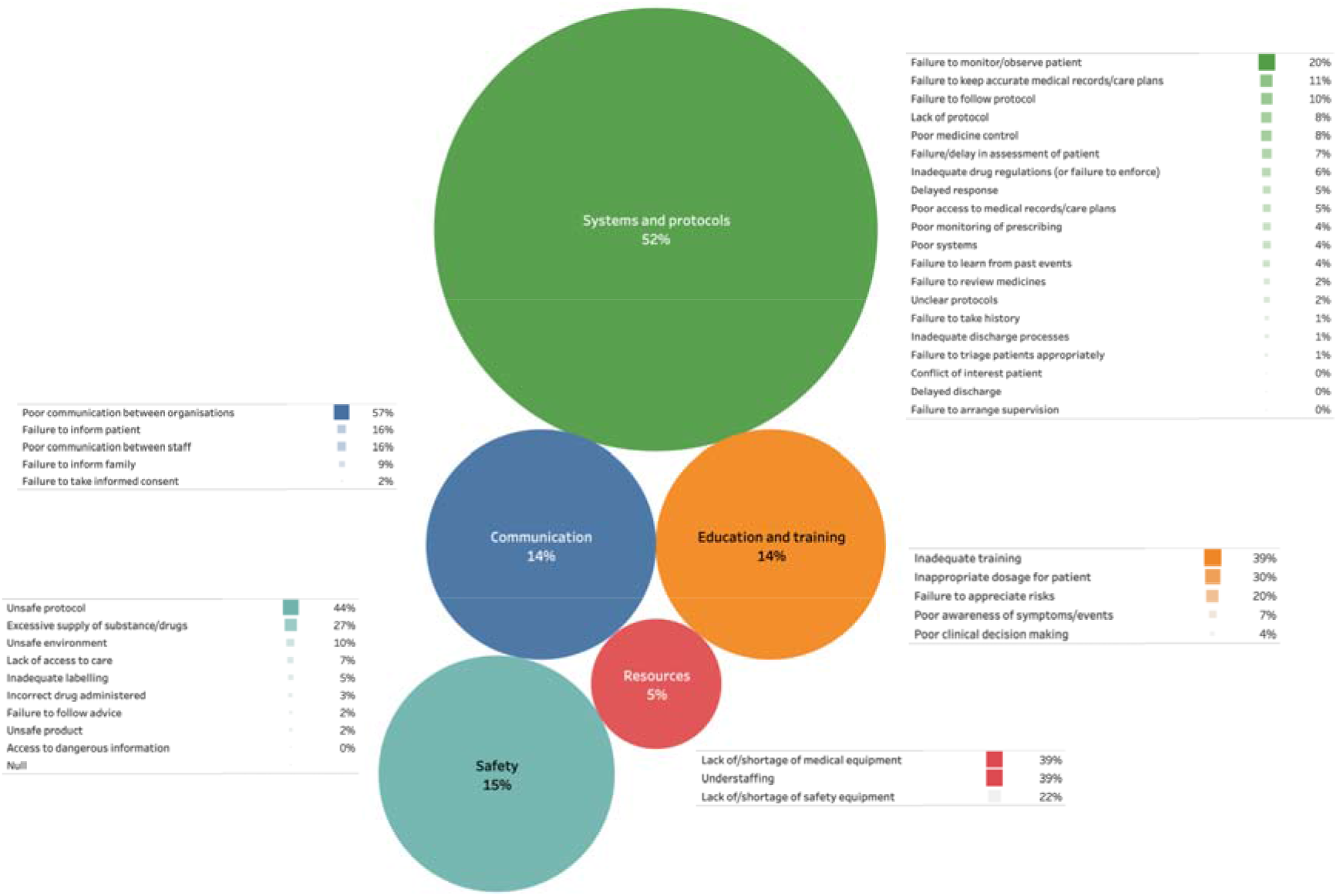
Concerns raised by coroners in 219 opioid-related Prevention of Future Deaths reports (PFDs) in England and Wales published between July 2013 and February 2022. The size of each circle is representative of the frequency of the fiver higher order themes. The individual concerns and their frequencies are reported in adjacent lists.

Coroners sent 360 PFDs to 43 unique organisations (Table 1). The National Health Service (NHS) in England and Wales received the most (51%) PFDs; however most (53%) of their responses were overdue (i.e. no response within 56 days of receiving the report). The response rate varied widely between organisations: 36% responded early or on time, 11% responded late, and 53% remained overdue as of 23 February 2022.

**Table 1.**
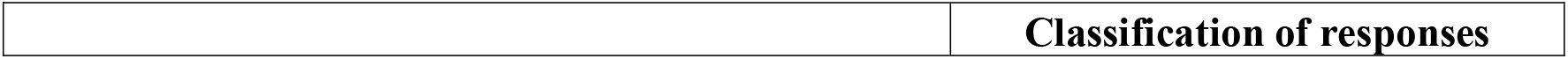
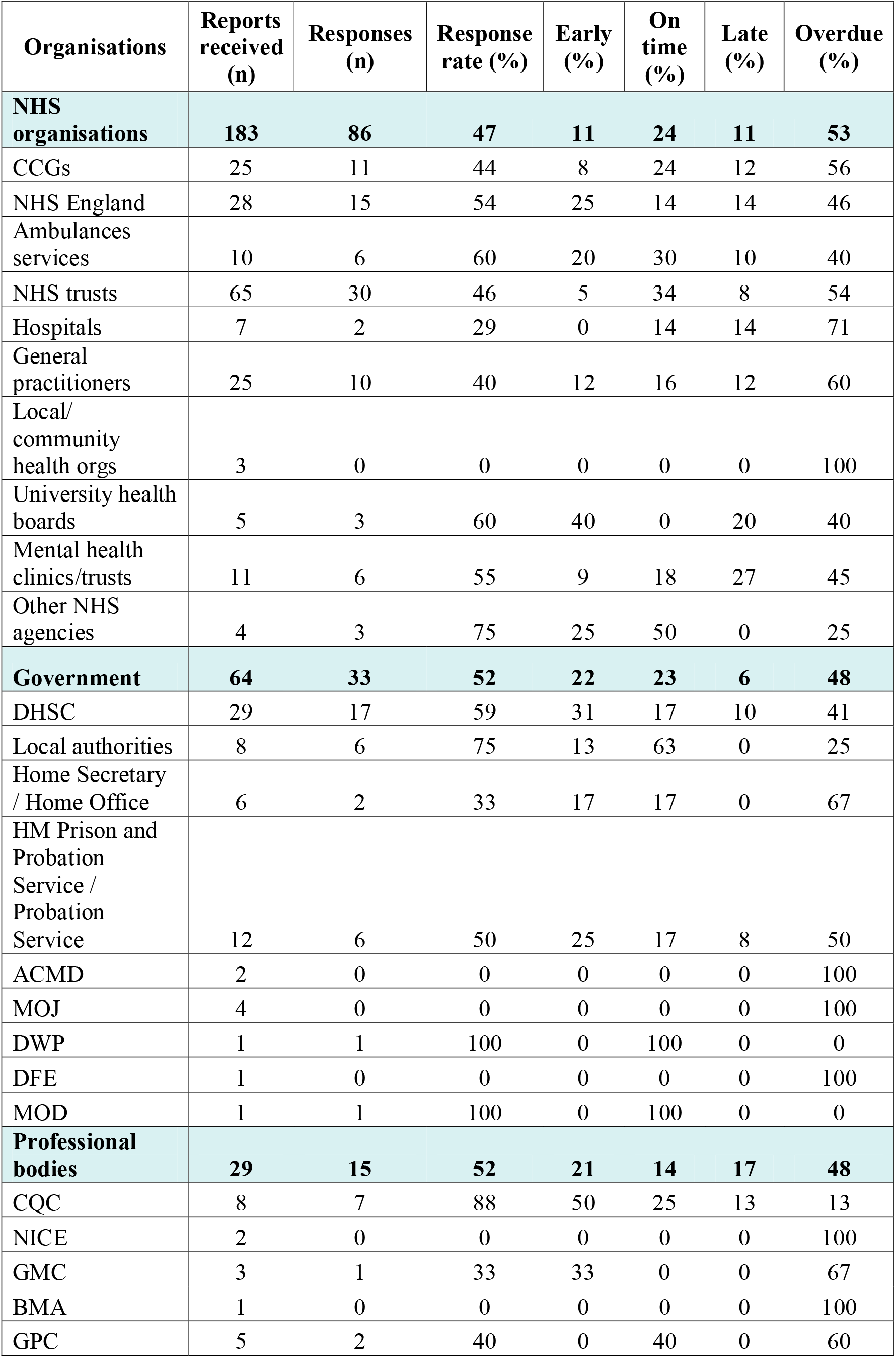
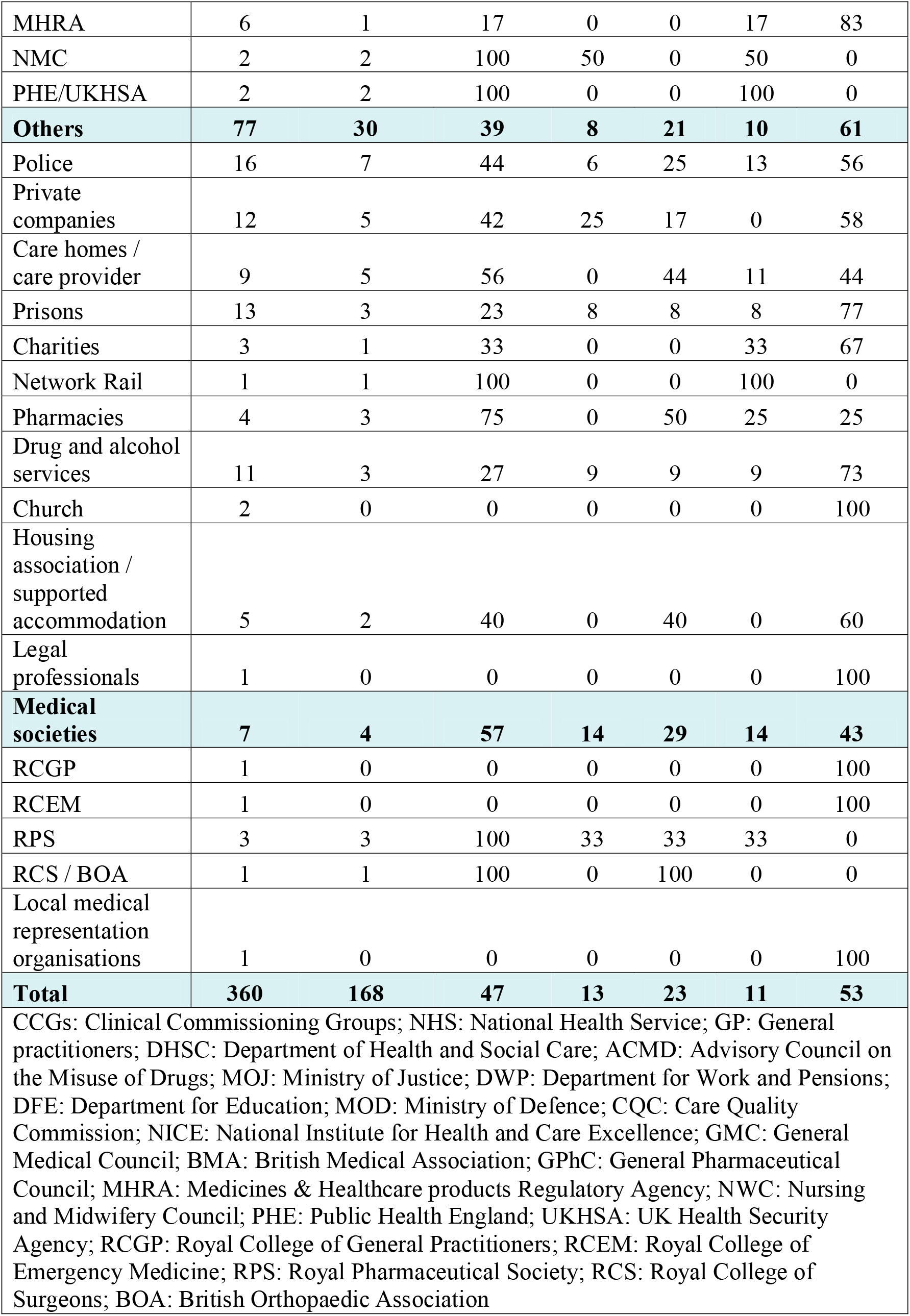
Organisations and individuals who received the 219 opioid-related Prevention of Future Deaths reports (PFDs) in England and Wales published between July 2013 and February 2022 and their response rates according to Regulation 29 of the Coroners (Investigations) Regulations 2013. Similar individual organisations have been grouped together for clarity, e.g pharmacies.

| **Organisations** | **Reports received (n)** | **Responses (n)** | **Response rate (%)** | **Early (%)** | **On time (%)** | **Late (%)** | **Overdue (%)** |
| --- | --- | --- | --- | --- | --- | --- | --- |
| **NHS organisations** | **183** | **86** | **47** | **11** | **24** | **11** | **53** |
| CCGs | 25 | 11 | 44 | 8 | 24 | 12 | 56 |
| NHS England | 28 | 15 | 54 | 25 | 14 | 14 | 46 |
| Ambulances services | 10 | 6 | 60 | 20 | 30 | 10 | 40 |
| NHS trusts | 65 | 30 | 46 | 5 | 34 | 8 | 54 |
| Hospitals | 7 | 2 | 29 | 0 | 14 | 14 | 71 |
| General practitioners | 25 | 10 | 40 | 12 | 16 | 12 | 60 |
| Local/ community health orgs | 3 | 0 | 0 | 0 | 0 | 0 | 100 |
| University health boards | 5 | 3 | 60 | 40 | 0 | 20 | 40 |
| Mental health clinics/trusts | 11 | 6 | 55 | 9 | 18 | 27 | 45 |
| Other NHS agencies | 4 | 3 | 75 | 25 | 50 | 0 | 25 |
| **Government** | **64** | **33** | **52** | **22** | **23** | **6** | **48** |
| DHSC | 29 | 17 | 59 | 31 | 17 | 10 | 41 |
| Local authorities | 8 | 6 | 75 | 13 | 63 | 0 | 25 |
| Home Secretary / Home Office | 6 | 2 | 33 | 17 | 17 | 0 | 67 |
| HM Prison and Probation Service / Probation Service | 12 | 6 | 50 | 25 | 17 | 8 | 50 |
| ACMD | 2 | 0 | 0 | 0 | 0 | 0 | 100 |
| MOJ | 4 | 0 | 0 | 0 | 0 | 0 | 100 |
| DWP | 1 | 1 | 100 | 0 | 100 | 0 | 0 |
| DFE | 1 | 0 | 0 | 0 | 0 | 0 | 100 |
| MOD | 1 | 1 | 100 | 0 | 100 | 0 | 0 |
| **Professional bodies** | **29** | **15** | **52** | **21** | **14** | **17** | **48** |
| CQC | 8 | 7 | 88 | 50 | 25 | 13 | 13 |
| NICE | 2 | 0 | 0 | 0 | 0 | 0 | 100 |
| GMC | 3 | 1 | 33 | 33 | 0 | 0 | 67 |
| BMA | 1 | 0 | 0 | 0 | 0 | 0 | 100 |
| GPC | 5 | 2 | 40 | 0 | 40 | 0 | 60 |
| MHRA | 6 | 1 | 17 | 0 | 0 | 17 | 83 |
| NMC | 2 | 2 | 100 | 50 | 0 | 50 | 0 |
| PHE/UKHSA | 2 | 2 | 100 | 0 | 0 | 100 | 0 |
| **Others** | **77** | **30** | **39** | **8** | **21** | **10** | **61** |
| Police | 16 | 7 | 44 | 6 | 25 | 13 | 56 |
| Private companies | 12 | 5 | 42 | 25 | 17 | 0 | 58 |
| Care homes / care provider | 9 | 5 | 56 | 0 | 44 | 11 | 44 |
| Prisons | 13 | 3 | 23 | 8 | 8 | 8 | 77 |
| Charities | 3 | 1 | 33 | 0 | 0 | 33 | 67 |
| Network Rail | 1 | 1 | 100 | 0 | 0 | 100 | 0 |
| Pharmacies | 4 | 3 | 75 | 0 | 50 | 25 | 25 |
| Drug and alcohol services | 11 | 3 | 27 | 9 | 9 | 9 | 73 |
| Church | 2 | 0 | 0 | 0 | 0 | 0 | 100 |
| Housing association / supported accommodation | 5 | 2 | 40 | 0 | 40 | 0 | 60 |
| Legal professionals | 1 | 0 | 0 | 0 | 0 | 0 | 100 |
| **Medical societies** | **7** | **4** | **57** | **14** | **29** | **14** | **43** |
| RCGP | 1 | 0 | 0 | 0 | 0 | 0 | 100 |
| RCEM | 1 | 0 | 0 | 0 | 0 | 0 | 100 |
| RPS | 3 | 3 | 100 | 33 | 33 | 33 | 0 |
| RCS / BOA | 1 | 1 | 100 | 0 | 100 | 0 | 0 |
| Local medical representation organisations | 1 | 0 | 0 | 0 | 0 | 0 | 100 |
| **Total** | **360** | **168** | **47** | **13** | **23** | **11** | **53** |
| CCGs: Clinical Commissioning Groups; NHS: National Health Service; GP: General practitioners; DHSC: Department of Health and Social Care; ACMD: Advisory Council on the Misuse of Drugs; MOJ: Ministry of Justice; DWP: Department for Work and Pensions; DFE: Department for Education; MOD: Ministry of Defence; CQC: Care Quality Commission; NICE: National Institute for Health and Care Excellence; GMC: General Medical Council; BMA: British Medical Association; GPhC: General Pharmaceutical Council; MHRA: Medicines & Healthcare products Regulatory Agency; NWC: Nursing and Midwifery Council; PHE: Public Health England; UKHSA: UK Health Security Agency; RCGP: Royal College of General Practitioners; RCEM: Royal College of Emergency Medicine; RPS: Royal Pharmaceutical Society; RCS: Royal College of Surgeons; BOA: British Orthopaedic Association |  |  |  |  |  |  |  |

### Classification of responses

CCGs: Clinical Commissioning Groups; NHS: National Health Service; GP: General practitioners; DHSC: Department of Health and Social Care; ACMD: Advisory Council on the Misuse of Drugs; MOJ: Ministry of Justice; DWP: Department for Work and Pensions; DFE: Department for Education; MOD: Ministry of Defence; CQC: Care Quality Commission; NICE: National Institute for Health and Care Excellence; GMC: General Medical Council; BMA: British Medical Association; GPhC: General Pharmaceutical Council; MHRA: Medicines & Healthcare products Regulatory Agency; NWC: Nursing and Midwifery Council; PHE: Public Health England; UKHSA: UK Health Security Agency; RCGP: Royal College of General Practitioners; RCEM: Royal College of Emergency Medicine; RPS: Royal Pharmaceutical Society; RCS: Royal College of Surgeons; BOA: British Orthopaedic Association

## Discussion

There were 219 deaths involving opioids where coroners believed actions should be taken to prevent similar deaths. On average, 32 years of life were lost for every opioid-related PFD. Morphine, methadone, and heroin were responsible for most deaths, which frequently occurred in London, the North-West, and the South-West of England. Coroners raised repeated concerns relating to systems and protocols, such as failures to arrange appropriate monitoring or follow-up, and the maintenance of accurate medical records and plans of patients receiving opioids. These concerns were sent to hundreds of organisations, mostly healthcare providers, and NHS services. Despite the legal requirements to do so, less than half of all recipients have responded, or have had their responses published in the public domain. It is therefore unclear what actions are being taken to prevent avoidable opioid-related deaths.

### Comparison with existing literature

Several studies have investigated coroners’ concerns in PFDs^9-14^, yet none has focused exclusively on opioid-related deaths, although opioids have previously been implicated as the most common medicine type causing preventable deaths^14^. These studies have similarly found poor response rates from recipients receiving PFDs^9,25^ and wide geographical variation in the writing of reports^9,26^, concerns that were also highlighted in a 2021 report by the House of Commons Justice Select Committee.^27^ A case series of anticoagulant-related PFDs similarly showed that systems and protocols were the most frequent coroners’ concerns.^9^

Several of our findings relating to opioid-related deaths have been reflected in the literature. An observational study of 76 countries published in 2021 showed that the UK had the highest rate of opioid consumption, which was driven by a greater use of tramadol and codeine in the UK^1^ compared with other countries. In line with this research, we found that tramadol and codeine were the fourth and fifth most common opioids implicated in deaths. An analysis of over-the-counter sales of codeine in 31 countries also showed that the UK had the fourth highest rates of sales,^28^ which may have contributed to the one in ten codeine-related PFD deaths we identified. A retrospective cohort study of adults in Glasgow, Scotland, showed that the co-occurrence of opioid dependence with homelessness, involvement with criminal justice services, and psychosis was associated with higher rates of premature mortality.^29^ Similarly, we found that 14% of the PFDs in our analysis were categorised on the Courts and Tribunals Judiciary website as ‘Mental health related deaths’ and 13% as ‘State Custody’ and ‘Police related deaths’. A matched cohort analysis of causes of deaths of English people who used illicit opioids showed that fatal poisonings were more common among younger individuals, whereas older individuals were more likely to die from non-communicable diseases.^30^ This may explain the lower average age of death among those who sourced opioids illicitly in PFDs. A case cross-over study of opioid-related deaths showed that while the prescription of high-dose opioids or co-prescription of gabapentinoids or antidepressants increased the risk of death, considerable proportions of individuals had not been given an opioid in the year prior to death (26·7%) or had been given relatively low doses (22%).^31^ This suggests that a considerable number of deaths may have been accidental overdoses, as a result of illicit use or use of stockpiled prescribed drugs, both of which are scenarios found in several of the cases in our analysis.

### Strengths and limitations

Our study builds on prior research that examined samples and subgroups of PFDs.^11,25,26^ Using reproducible data collection methods, we efficiently collected all available PFDs for this study.^17^ However, the 219 opioid-related PFDs identified cannot represent all preventable deaths involving opioids in England and Wales as coronial variation in the writing of PFDs has been reported.^27,32^ The issuing of PFDs also depends on the working practices of individual coroners, who, although required to submit PFDs when they consider that actions should be taken to prevent further deaths, are not subject to auditing or quality assurance of the reports they write. We controlled for this variation by reporting opioid-related PFDs as a proportion of all PFDs and the rates for each geographical region. We also identified missing information in the PFDs; for example, 37% of PFDs did not include age at death or date of birth. This could be improved by standardising the reporting of information in PFDs using electronic forms and providing PFD training for coroners.

### Implications for practice and policy

Understanding causes of death and how they can be prevented is critical for improving health, economic and social outcomes globally. Although the rate of opioid-related deaths differs between North America and the UK,^33^ there are calls for the UK Government to recognise the rise in opioid-related deaths as a public health crisis.^34,35^ Both Public Health England (PHE) and the British Medical Association (BMA) have released reports that identified the widespread use and harms of opioids with recommendations to tackle these issues,^36,37^ but these recommendations have largely remained unimplemented.^36,37^ Our findings add to these reports by providing insight into the causes of deaths involving opioids that can be prevented. NHS England report that they review PFDs to develop guidance and advice.^38^ However, it is not clear how the insights from PFDs are being used in practice and policy.

Studies suggest that practitioners will engage with safety concerns relating to opioid deaths. A randomised trial of clinicians prescribing opioids to patients in the USA who subsequently died from overdose showed that those who received notification of their patients’ deaths reduced their prescribing of opioids, initiated fewer prescriptions, and prescribed fewer high-dose opioids, compared with clinicians who were provided with a safe prescribing injunction or no intervention.^39^ Such feedback-based approaches for opioids have been shown to be similarly effective in primary care settings in the UK.^40^ However, further research of the most effective and safe way to reduce opioid-related deaths stills requires investigation.

Problems with the PFD system^27^, a lack of functional access to reports, and poor response rates and dissemination may be hindering the national implementation of findings from PFDs. Future research should explore the barriers to and facilitators of writing and responding to PFDs and assess the effectiveness of PFDs in changing clinical practice and policy, and in reducing preventable deaths. An analysis of the 168 responses submitted by organisations who received opioid-related PFDs is also required, to identify the actions taken and proposed to prevent deaths, as well as the gaps that have not been addressed.

## Conclusions

In conclusion, we have used reproducible data-collection methods to comprehensively analyse all available PFDs, which provide valuable lessons for preventing future opioid-related deaths. The dissemination of these lessons remains limited, but implementation of their findings in future guidelines and clinical practice will improve patient safety and reduce the years of life lost from opioids-related deaths.

## Data Availability

All study materials, data, and statistical code are openly available via online repositories. The study protocol was preregistered on the Open Science Framework (OSF; https://doi.org/10.17605/OSF.IO/QJE8A); the code to generate the database and the Preventable Deaths Tracker is openly available via GitHub (https://github.com/georgiarichards/georgiarichards.github.io); individual Prevention of Future Deaths reports are available on the Courts and Tribunals Judiciary website (https://www.judiciary.uk/prevention-of-future-death-reports/); all other study materials are openly available via the OSF project page (https://doi.org/10.17605/OSF.IO/ECZ4R).

## Authors and contributors

FD updated the study protocol, finalised the screening of PFDs, extracted remaining data, conducted all analyses, produced all figures and tables, and wrote the first draft of the manuscript. GCR conceptualised, designed, and initiated the study; provided supervisory support, and edited the first draft of the manuscript. MB and HSF assisted with screening PFDs for inclusion. ET assisted with data extraction. NJD wrote and supervised the initial code for the web scraper to collect data. REF and ARC contributed to study conceptualisation. CH and JKA provided supervisory support and oversight. All study authors read, contributed to, and approved the final manuscript.

