## Supplementary Figure S1 for "Preventable deaths involving opioids in England and Wales, 2013-2022: a systematic case series of coroners’ reports"

**Supplementary Figure S1.** Algorithm for case inclusion

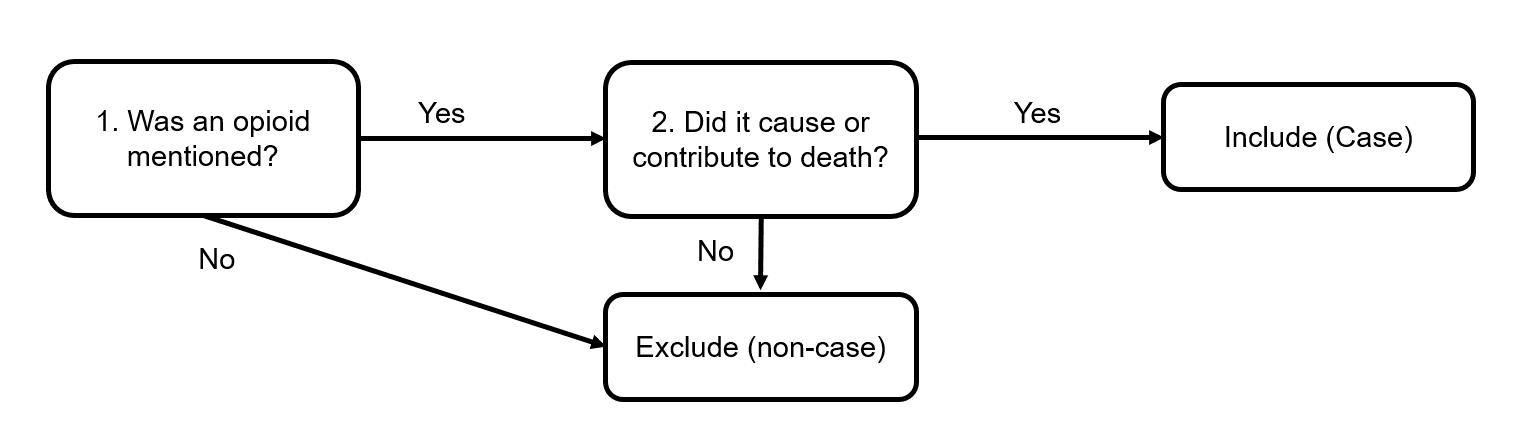

**Supplementary Table S1.** Opioid-related PFDs and deaths each year, from July 2013 to February 2022

| **Year** | **Total number of PFDs** | **Number of opioid-related PFDs** | **Rate of opioid-related PFDs** | **Number of ONS opioid deaths** | **Percentage of opioid PFDs vs ONS opioid deaths** |
| --- | --- | --- | --- | --- | --- |
| 2013 | 178 | 4 | 2.25% | 1884 | n/a* |
| 2014 | 564 | 28 | 4.96% | 2108 | 1.33% |
| 2015 | 489 | 23 | 4.70% | 2377 | 0.97% |
| 2016 | 476 | 17 | 3.57% | 2395 | 0.71% |
| 2017 | 445 | 29 | 6.52% | 2355 | 1.23% |
| 2018 | 420 | 33 | 7.86% | 2644 | 1.25% |
| 2019 | 527 | 43 | 8.16% | 2568 | 1.67% |
| 2020 | 312 | 18 | 5.77% | 2730 | 0.66% |
| 2021 | 424 | 21 | 4.95% | n/a† | n/a |
| 2022 | 41 | 3 | 7.32% | n/a† | n/a |
| **Total** | **3876** | **219** | **5.65%** | **19061** | **1.15%** |
| **Median (IQR)** | 460.5 (423-498.5) | 25.5 (20.25-30) | 5.37% (4.89-6.85%) | 2386 (2293.25-2587) | 1.23% (0.84-1.29%) |
| * Data only from July 2013 onwards  † ONS figures for opioid deaths only up to 2020 | | | | | |

**Supplementary Table S2.** Baseline characteristics of each sub-group

|  | **Illicit** | **Prescribed** | **Combination** |
| --- | --- | --- | --- |
| **Number** | 52 | 114 | 31 |
| **Age** | 36.9 | 46.3 | 41 |
| **% Male** | 86.5 | 50 | 77.4 |

**Supplementary Table S3.** Categorisation of 219 Prevention of Future Deaths reports (PFDs) as per their categories on the judiciary website. Note that individual PFDs may be tagged with multiple categories.

| **Category** | **Number** |
| --- | --- |
| Alcohol, drug and medication related deaths | 82 |
| Hospital Death | 80 |
| Community Healthcare | 56 |
| Emergency Services Related Death | 37 |
| Mental health related deaths | 30 |
| State Custody related deaths | 22 |
| Other related deaths | 20 |
| Suicide (from 2015) | 15 |
| Product related deaths | 8 |
| Police related deaths | 6 |
| Care home related deaths | 4 |
| Child death (from 2015) | 2 |
| Railway related deaths | 1 |
| Wales (from 2019) | 1 |

**Supplementary Table S4**. Causes of death within opioid-related PFDs, by ICD-10 criteria. Note that coding of an individual death can involve multiple ICD-10 codes. ICD-10 codes with a prevalence of >1% are highlighted in green.

| **ICD-10 Code** | **Number** | **Percentage** | **Specific** |
| --- | --- | --- | --- |
| T40.2 | 89 | 40.64 | Poisoning by 'other opioids' (codeine, morphine) |
| T40.3 | 43 | 19.63 | Poisoning by methadone |
| T40.4 | 41 | 18.72 | Poisoning by other synthetic narcotics |
| T40.1 | 30 | 13.70 | Poisoning by heroin |
| T42.4 | 26 | 11.87 | Poisoning by benzodiazepines |
| T43.8 | 20 | 9.13 | Poisoning by other psychotropic drugs, not elsewhere specified |
| T51.0 | 20 | 9.13 | Toxic effect of alcohol |
| T43.2 | 15 | 6.85 | Poisoning by other and unspecified antidepressants |
| T43.0 | 14 | 6.39 | Poisoning by TCA |
| T40 | 11 | 5.02 | Poisoning by narcotics and psychodysleptics [hallucinogens] |
| T40.5 | 10 | 4.57 | Poisoning by cocaine |
| T42.6 | 9 | 4.11 | Poisoning by other antiepileptic and sedative-hypnotic drugs |
| J69.0 | 7 | 3.20 | Pneumonitis/aspiration |
| T39.1 | 7 | 3.20 | Poisoning: 4-Aminophenol derivatives |
| J18.0 | 6 | 2.74 | Bronchopneumonia |
| R09.2 | 3 | 1.37 | Respiratory arrest |
| T40.7 | 3 | 1.37 | Poisoning by cannabis (derivatives) |
| G93.1 | 2 | 0.91 |  |
| I26 | 2 | 0.91 |  |
| I46 | 2 | 0.91 |  |
| J96.0 | 2 | 0.91 |  |
| T17 | 2 | 0.91 |  |
| T39.3 | 2 | 0.91 |  |
| T43.5 | 2 | 0.91 |  |
| T44.7 | 2 | 0.91 |  |
| T47.6 | 2 | 0.91 |  |
| T68 | 2 | 0.91 |  |
| A40.3 | 1 | 0.46 |  |
| C34 | 1 | 0.46 |  |
| C78 | 1 | 0.46 |  |
| D68.3 | 1 | 0.46 |  |
| I21.9 | 1 | 0.46 |  |
| I25.1 | 1 | 0.46 |  |
| I50.0 | 1 | 0.46 |  |
| I50.1 | 1 | 0.46 |  |
| J18.9 | 1 | 0.46 |  |
| J81 | 1 | 0.46 |  |
| K25 | 1 | 0.46 |  |
| K55.0 | 1 | 0.46 |  |
| K63.1 | 1 | 0.46 |  |
| K85 | 1 | 0.46 |  |
| K91.8 | 1 | 0.46 |  |
| K92.2 | 1 | 0.46 |  |
| L89.3 | 1 | 0.46 |  |
| P29.1 | 1 | 0.46 |  |
| S22.3 | 1 | 0.46 |  |
| T01.8 | 1 | 0.46 |  |
| T36.1 | 1 | 0.46 |  |
| T41.2 | 1 | 0.46 |  |
| T41.3 | 1 | 0.46 |  |
| T43.3 | 1 | 0.46 |  |
| T43.6 | 1 | 0.46 |  |
| T45.0 | 1 | 0.46 |  |
| T48.3 | 1 | 0.46 |  |
| T79.6 | 1 | 0.46 |  |
| TO1.8 | 1 | 0.46 |  |
| W01 | 1 | 0.46 |  |
| X71 | 1 | 0.46 |  |
| X78 | 1 | 0.46 |  |
| Y55.1 | 1 | 0.46 |  |

**Supplementary Figure S2.** Frequency of different opioid drugs implicated in PFDs for each subgroup

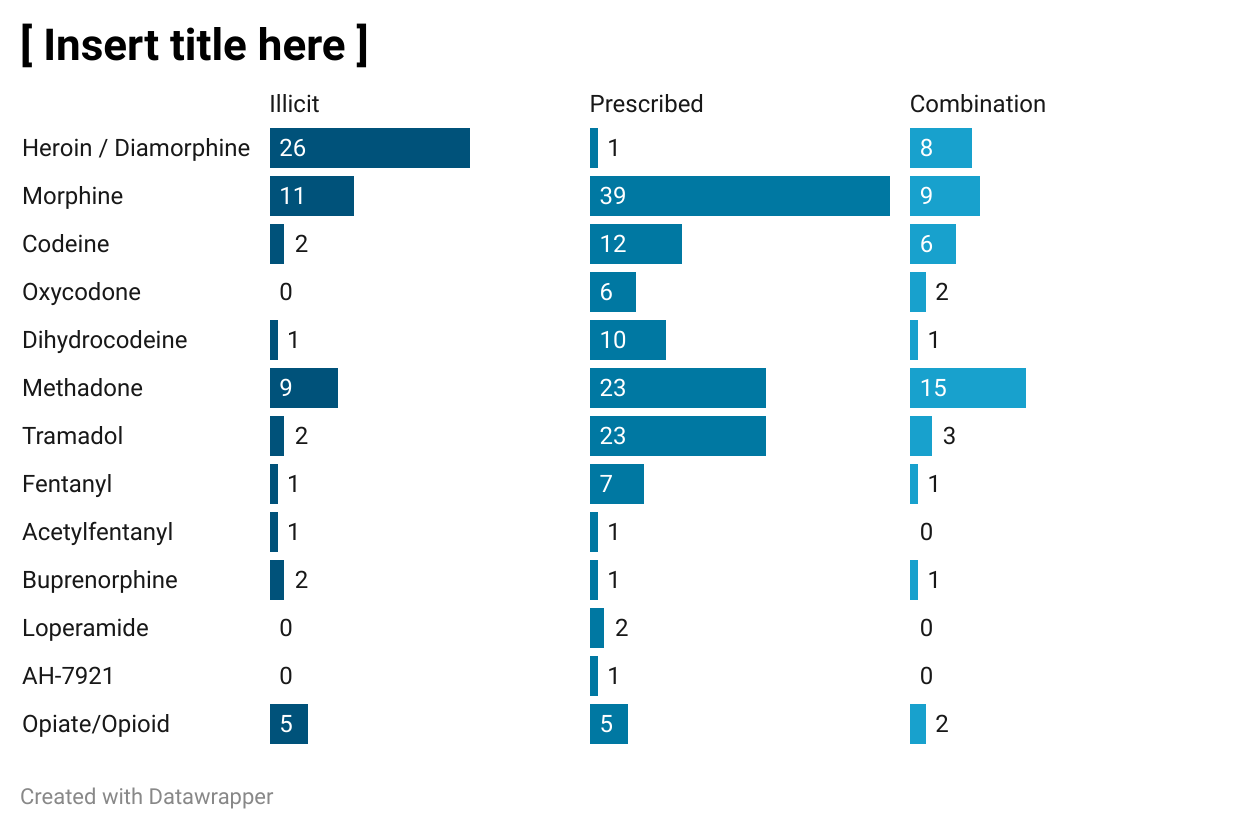

**Supplementary Table S5.** Coroner areas and number of opioid-related PFDs published between July 2013 and February 2022. Note that during the time period of the study, various coroner areas have undergone mergers. The coroner area names reported here are those written on the PFD at the time of the PFD’s publication.

| **Coroner Area** | **Number of PFDs** |
| --- | --- |
| Avon | 4 |
| Bedfordshire & Luton | 4 |
| Berkshire | 2 |
| Birmingham and Solihull | 7 |
| Black Country | 6 |
| Blackburn, Hyndburn & Ribble Valley | 1 |
| Blackpool & Fylde | 4 |
| Brighton and Hove | 4 |
| Buckinghamshire | 1 |
| Cambridgeshire & Peterborough | 1 |
| Cardiff & the Vale of Glamorgan | 2 |
| Cheshire | 1 |
| Cornwall & the Isle of Scilly | 7 |
| County Durham & Darlington | 5 |
| Coventry | 2 |
| Cumbria | 2 |
| Cumbria (South & East) | 1 |
| Derby and Derbyshire | 2 |
| Dorset | 4 |
| East Riding and Kingston Upon Hull | 1 |
| East Sussex | 1 |
| Essex | 6 |
| Exeter & Greater Devon | 6 |
| Gateshead & South Tyneside | 1 |
| Gloucestershire | 4 |
| Greater Manchester South | 4 |
| Gwent | 1 |
| Hampshire (Central) | 1 |
| Hertfordshire | 2 |
| Isle of Wight | 1 |
| Lancashire and Blackburn with Darwen | 3 |
| Leicester City & South Leicestershire | 1 |
| Lincolnshire (Central) | 1 |
| Liverpool | 2 |
| London | 1 |
| London (East) | 3 |
| London (North) | 1 |
| London (South) | 1 |
| London (West) | 4 |
| London Inner (North) | 9 |
| London Inner (South) | 7 |
| London Inner (West) | 8 |
| Manchester (City) | 4 |
| Manchester (North) | 10 |
| Manchester (South) | 18 |
| Manchester (West) | 5 |
| Milton Keynes | 1 |
| Norfolk | 5 |
| North East Kent | 1 |
| North Wales (East & Central) | 2 |
| North West Kent | 1 |
| Nottinghamshire | 3 |
| Plymouth Torbay and South Devon | 1 |
| Portsmouth & South East Hampshire | 1 |
| Shropshire, Telford & Wrekin | 1 |
| Somerset (West) | 1 |
| South Wales Central | 3 |
| South Yorkshire (East) | 3 |
| South Yorkshire (West) | 2 |
| Southampton and New Forest | 1 |
| Staffordshire South | 3 |
| Stoke-on-Trent & North Staffordshire | 3 |
| Suffolk | 3 |
| Surrey | 1 |
| Swansea, Neath and Port Talbot | 1 |
| West Sussex | 4 |
| West Yorkshire (East) | 2 |
| West Yorkshire (West) | 2 |
| Wiltshire & Swindon | 5 |
| Worcestershire | 2 |
| York | 1 |

**Supplementary table S6.** Concerns raised by coroners grouped by higher-order theme following content analysis

| **Themes (% of concerns)** | **Concern** | **Cases, n (%)** |
| --- | --- | --- |
| **Systems and protocols** | failure to monitor/observe patient | 41 (19) |
|  | failure to keep accurate medical records/care plans | 23 (11) |
|  | failure to follow protocol | 20 (9) |
|  | poor medicine control | 17 (8) |
|  | lack of protocol | 17 (8) |
|  | failure/delay in assessment of patient | 14 (6) |
|  | inadequate drug regulations (or failure to enforce) | 12 (5) |
|  | delayed response | 10 (5) |
|  | poor access to medical records/care plans | 10 (5) |
|  | poor monitoring of prescribing | 9 (4) |
|  | poor systems | 9 (4) |
|  | failure to learn from past events | 8 (4) |
|  | unclear protocols | 5 (2) |
|  | failure to review medicines | 5 (2) |
|  | inadequate discharge processes | 3 (1) |
|  | failure to take history | 3 (1) |
|  | failure to triage patients appropriately | 2 (1) |
|  | failure to arrange supervision | 0 (0) |
|  | Delayed discharge | 0 (0) |
|  | Conflict of interest patient | 0 (0) |
| **Safety** | unsafe protocol | 26 (12) |
|  | excessive supply of substance/drugs | 16 (7) |
|  | unsafe environment | 6 (3) |
|  | lack of access to care | 4 (2) |
|  | inadequate labelling | 3 (1) |
|  | incorrect drug administered | 2 (1) |
|  | unsafe product | 1 (0) |
|  | failure to follow advice | 1 (0) |
|  | Access to dangerous information | 0 (0) |
| **Communication** | poor communication between organisations | 32 (15) |
|  | poor communication between staff | 9 (4) |
|  | failure to inform patient | 9 (4) |
|  | failure to inform family | 5 (2) |
|  | failure to take informed consent | 1 (0) |
| **Education and training** | inadequate training | 22 (10) |
|  | inappropriate dosage for patient | 17 (8) |
|  | failure to appreciate risks | 11 (5) |
|  | poor awareness of symptoms/events | 4 (2) |
|  | poor clinical decision making | 2 (1) |
| **Resources** | understaffing | 7 (3) |
|  | lack of/shortage of medical equipment | 7 (3) |
|  | lack of/shortage of safety equipment | 4 (2) |

**Supplementary Table S7**. Concerns raised by coroners grouped by higher-order theme following content analysis, and split by sub-group. Concerns raised in more than 10% of the PFDs within a sub-group are highlighted in yellow.

|  |  | **Illicit** | **Prescribed** | **Combination** |
| --- | --- | --- | --- | --- |
| **Systems and protocols** | unclear protocols | 3 (6) | 2 (2) | 0 (0) |
|  | poor medicine control | 6 (12) | 7 (6) | 4 (13) |
|  | poor monitoring of prescribing | 2 (4) | 7 (6) | 0 (0) |
|  | poor systems | 2 (4) | 2 (2) | 2 (6) |
|  | lack of protocol | 2 (4) | 11 (10) | 2 (6) |
|  | inadequate drug regulations (or failure to enforce) | 5 (10) | 5 (4) | 2 (6) |
|  | inadequate discharge processes | 0 (0) | 1 (1) | 1 (3) |
|  | failure/delay in assessment of patient | 3 (6) | 6 (5) | 2 (6) |
|  | failure to triage patients appropriately | 0 (0) | 2 (2) | 0 (0) |
|  | delayed response | 1 (2) | 5 (4) | 2 (6) |
|  | failure to follow protocol | 4 (8) | 11 (10) | 3 (10) |
|  | failure to keep accurate medical records/care plans | 3 (6) | 13 (11) | 5 (16) |
|  | failure to learn from past events | 1 (2) | 6 (5) | 1 (3) |
|  | failure to arrange supervision | 0 (0) | 0 (0) | 0 (0) |
|  | failure to monitor/observe patient | 11 (21) | 17 (15) | 10 (32) |
|  | failure to review medicines | 1 (2) | 4 (4) | 0 (0) |
|  | failure to take history | 1 (2) | 1 (1) | 1 (3) |
|  | poor access to medical records/care plans | 1 (2) | 7 (6) | 2 (6) |
|  | Delayed discharge | 0 (0) | 0 (0) | 0 (0) |
|  | Conflict of interest patient | 0 (0) | 0 (0) | 0 (0) |
| **Communication** | poor communication between organisations | 3 (6) | 18 (16) | 7 (23) |
|  | poor communication between staff | 2 (4) | 4 (4) | 1 (3) |
|  | failure to inform family | 1 (2) | 3 (3) | 1 (3) |
|  | failure to inform patient | 3 (6) | 5 (4) | 0 (0) |
|  | failure to take informed consent | 1 (2) | 0 (0) | 0 (0) |
| **Safety** | Access to dangerous information | 0 (0) | 0 (0) | 0 (0) |
|  | unsafe environment | 4 (8) | 2 (2) | 0 (0) |
|  | unsafe protocol | 7 (13) | 12 (11) | 5 (16) |
|  | unsafe product | 0 (0) | 0 (0) | 0 (0) |
|  | lack of access to care | 1 (2) | 2 (2) | 1 (3) |
|  | incorrect drug administered | 0 (0) | 1 (1) | 1 (3) |
|  | inadequate labelling | 0 (0) | 3 (3) | 0 (0) |
|  | excessive supply of substance/drugs | 1 (2) | 15 (13) | 0 (0) |
|  | failure to follow advice | 0 (0) | 0 (0) | 1 (3) |
| **Education and training** | inappropriate dosage for patient | 1 (2) | 14 (12) | 2 (6) |
|  | inadequate training | 7 (13) | 11 (10) | 1 (3) |
|  | failure to appreciate risks | 3 (6) | 3 (3) | 3 (10) |
|  | poor awareness of symptoms/events | 1 (2) | 3 (3) | 0 (0) |
|  | poor clinical decision making | 0 (0) | 2 (2) | 0 (0) |
| **Resources** | understaffing | 2 (4) | 1 (1) | 2 (6) |
|  | lack of/shortage of safety equipment | 2 (4) | 1 (1) | 1 (3) |
|  | lack of/shortage of medical equipment | 1 (2) | 3 (3) | 0 (0) |
